# Multiple SARS-CoV-2 variants escape neutralization by vaccine-induced humoral immunity

**DOI:** 10.1101/2021.02.14.21251704

**Authors:** Wilfredo F. Garcia-Beltran, Evan C. Lam, Kerri St. Denis, Adam D. Nitido, Zeidy H. Garcia, Blake M. Hauser, Jared Feldman, Maia N. Pavlovic, David J. Gregory, Mark C. Poznansky, Alex Sigal, Aaron G. Schmidt, A. John Iafrate, Vivek Naranbhai, Alejandro B. Balazs

## Abstract

Vaccination elicits immune responses capable of potently neutralizing SARS-CoV-2. However, ongoing surveillance has revealed the emergence of variants harboring mutations in spike, the main target of neutralizing antibodies. To understand the impact of these variants, we evaluated the neutralization potency of 99 individuals that received one or two doses of either BNT162b2 or mRNA-1273 vaccines against pseudoviruses representing 10 globally circulating strains of SARS-CoV-2. Five of the 10 pseudoviruses, harboring receptor-binding domain mutations, including K417N/T, E484K, and N501Y, were highly resistant to neutralization. Cross-neutralization of B.1.351 variants was comparable to SARS-CoV and bat-derived WIV1-CoV, suggesting that a relatively small number of mutations can mediate potent escape from vaccine responses. While the clinical impact of neutralization resistance remains uncertain, these results highlight the potential for variants to escape from neutralizing humoral immunity and emphasize the need to develop broadly protective interventions against the evolving pandemic.

## INTRODUCTION

Since the first described human infection with severe acute respiratory syndrome coronavirus 2 (SARS-CoV-2) in December of 2019, nine vaccines have been approved for use in humans (“COVID-19 Vaccine Tracker” n.d.). Two of the vaccines currently in use worldwide, BNT162b2 (manufactured by Pfizer) and mRNA-1273 (manufactured by Moderna), are based on lipid nanoparticle delivery of mRNA encoding a prefusion stabilized form of spike protein derived from SARS-CoV-2 isolated early in the epidemic from Wuhan, China. Both of these vaccines demonstrated >94% efficacy at preventing coronavirus disease 2019 (COVID-19) in phase III clinical studies performed in late 2020 in multiple countries (Polack et al. 2020; Baden et al. 2021). However, the recent emergence of novel circulating variants has raised significant concerns about geographic and temporal efficacy of these interventions. Indeed, more recently completed trials of two adenovirus-based vaccines (AZD1222 from Astrazeneca and JNJ-78436735 from Johnson & Johnson), a nanoparticle-based vaccine (NVX-CoV2373 from Novavax), and an inactivated protein vaccine (Coronavac) have demonstrated reduced overall efficacy (“Novavax COVID-19 Vaccine Demonstrates 89.3% Efficacy in UK Phase 3 Trial” n.d., “AZD1222 Vaccine Met Primary Efficacy Endpoint in Preventing COVID-19” 2020), and subset analyses suggest marked geographic variation with lower efficacy against mild-to-moderate disease in countries such as South Africa and Brazil where the epidemic is dominated by variant strains. Taken together, these data suggest that neutralization-resistant variants may have contributed to these outcomes (“South Africa Suspends Use of AstraZeneca’s COVID-19 Vaccine after It Fails to Clearly Stop Virus Variant” 2021; Herper et al. 2021).

One of the earliest variants that emerged and rapidly became globally dominant was D614G. While several studies demonstrated that this strain is more infectious (Korber et al. 2020; Yurkovetskiy et al. 2020a, [b] 2020; Plante et al. 2020; Zhou et al. 2020; Hou et al. 2020), we and others found that sera from convalescent individuals showed effective cross-neutralization of both wildtype and D614G variants (Garcia-Beltran et al. 2021; Legros et al. 2021; Hou et al. 2020). However, recent genomic surveillance in the United Kingdom has revealed rapid expansion of a novel lineage termed B.1.1.7 (also known as VOC-202012/01 or 501Y.V1). B.1.1.7 harbors 3 amino acid deletions and 7 missense mutations in spike, including D614G as well as N501Y in the ACE2 receptor-binding domain (RBD), and has been reported to be more infectious than D614G (Santos and Passos 2021; Galloway et al. 2021; Liu et al. 2021). Several studies have demonstrated that convalescent and vaccinee sera cross-neutralize B.1.1.7 variants with only slightly decreased potency, suggesting that prior infection or vaccination with wild-type SARS-CoV-2 may still provide protection against B.1.1.7 variants (Wu et al. 2021; Muik et al. 2021; Shen et al. 2021; Rees-Spear et al. 2021; P. Wang et al. 2021). There have also been reports of SARS-CoV-2 transmission between humans and minks in Denmark with a variant called mink cluster 5 or B.1.1.298, which harbors a 2-amino acid deletion and 4 missense mutations including Y453F in RBD. Concerns relating to ongoing interspecies transmission resulted in the culling of over 17 million Danish minks to prevent further viral spread and evolution (Oude Munnink et al. 2021; Oxner 2020). Another variant that recently emerged in California, United States, designated as B.1.429, contains 4 missense mutations in spike, one of which is a single L452R RBD mutation. The ability of B.1.1.298 and B.1.429 variants to evade neutralizing humoral immunity from prior infection or vaccination has yet to be determined.

Of particular concern is an E484K mutation in RBD, which was previously identified through *in vitro* selection experiments to escape from monoclonal antibodies (Baum et al. 2020) and was also recently identified through deep mutational scanning as a variant with the potential to evade monoclonal and serum antibody responses (Greaney et al. 2020, 2021). Novel variants arising from the B.1.1.28 lineage first described in Brazil and Japan, termed P.2 (with 3 spike missense mutations) and P.1 (with 12 spike missense mutations), contain this E484K mutation, and P.1 in particular also contains K417T and N501Y mutations in RBD. These strains have been spreading rapidly, and both P.2 and P.1 were recently found in documented cases of SARS-CoV-2 re-infection (Paiva et al. 2020; Faria et al. 2021; Resende et al. 2021; Naveca et al. 2021; Nonaka et al. 2021).

Of greatest concern has been the emergence of multiple strains of the B.1.351 lineage (also known as 501Y.V2), which were first reported in South Africa and have since spread globally (Tegally et al. 2021). This lineage bears three RBD mutations, K417N, E484K, and N501Y, in addition to several mutations outside of RBD, and several reports have suggested that convalescent and vaccinee sera have decreased cross-neutralization of B.1.351 lineage variants (P. Wang et al. 2021; Wibmer et al. 2021; Wu et al. 2021; Hu et al. 2021). A key limitation of several of these reports has been the use of single mutations or combinations of mutations that do not naturally occur. Regardless, the emergence of novel variants that appear to escape immune responses has spurred vaccine manufacturers to develop boosters for these spike variants (“Moderna Developing Booster Shot for New Virus Variant B.1.351” n.d.).

Here, we systematically assessed the neutralization potential of sera from a cohort of individuals who received one or two doses of the BNT162b2 (Pfizer) or mRNA-1273 (Moderna) vaccine against SARS-CoV-2 pseudoviruses that bear spike proteins found on circulating strains. We used our previously described high-throughput pseudovirus neutralization assay (Garcia-eltran et al. 2021) to quantify neutralization against variants first arising in the United Kingdom (B.1.1.7), Denmark (B.1.1.298), United States (B.1.429), Brazil and Japan (P.2 and P.1), and South Africa (three variants of the B.1.351 lineage), as well as SARS-CoV from the 2002 Hong Kong outbreak and the pre-emergent bat coronavirus WIV1-CoV. We find that although neutralization is largely preserved against many variants, those containing the K417N/T, E484K, and N501Y RBD mutations, namely, P.1 and B.1.351 variants, have significantly decreased neutralization even in fully vaccinated individuals. Individuals that received only a single recent dose of vaccine had weaker neutralization titers overall and did not exhibit detectable neutralization of B.1.351 variants in our assays.

Taken together, our results highlight that BNT162b2 and mRNA-1273 vaccines achieve only partial cross-neutralization of novel variants and support the reformulation of existing vaccines to include diverse spike sequences. Ultimately, development of new vaccines capable of eliciting broadly neutralizing antibodies may be necessary to resolve the ongoing pandemic.

## RESULTS

### Emergence and global spread of SARS-CoV-2 variants of concern

Since the first described case of SARS-CoV-2 infection in Wuhan, China in December of 2019, over 107 million confirmed infections have been documented (Coronavirus.jhu.edu), enabling viral diversification and the emergence of six distinct major lineages with numerous variants. A subset of these variants have been denoted as variants of concern by the World Health Organization given the presence of mutations with potential to increase transmissibility, virulence, or evade immune responses. We focused on variants of concern first described in the United Kingdom (B.1.1.7), Denmark (B.1.1.298), United States (B.1.429), Brazil and Japan (P.2 and P.1), and South Africa (B.1.351), most of which arose in late 2020 (**Figure 1A-B**). Although the naturally arising mutations in these variants span the entire spike protein, they mainly occur in S1 and RBD, the main target of neutralizing antibodies (**Figure 1C** and **S1**). After analyzing SARS-CoV-2 spike sequences deposited for each lineage in GISAID, we identified consensus sequence mutations that represent the dominant circulating strain for each lineage (**Figure 1D** and **S1**). In the case of the B.1.351 lineage, we studied three of the dominant variants comprising the majority of deposited sequences, which we designated v1, v2, and v3. The three main RBD mutations of concern are: (*i*) N501Y, present in B.1.1.7, P.1, and B.1.351 variants; (*ii*) E484K, present in the P.2, P.1, and B.1.351 variants; and (*iii*) K417T for the P.1 variant and K417N for the B.1.351 variants. Separately, the B.1.1.298 variant found in Danish minks contained a Y453F mutation in RBD, and the California variant B.1.429 contained an L452R. Although distantly related, SARS-CoV from the 2002 Hong Kong coronavirus outbreak and pre-emergent bat-derived WIV1-CoV both share ∼76% spike homology to SARS-CoV-2 and were assessed to provide comparison to serologically distinct coronaviruses (**Figure 1A** and **1D**).

**Figure 1.**
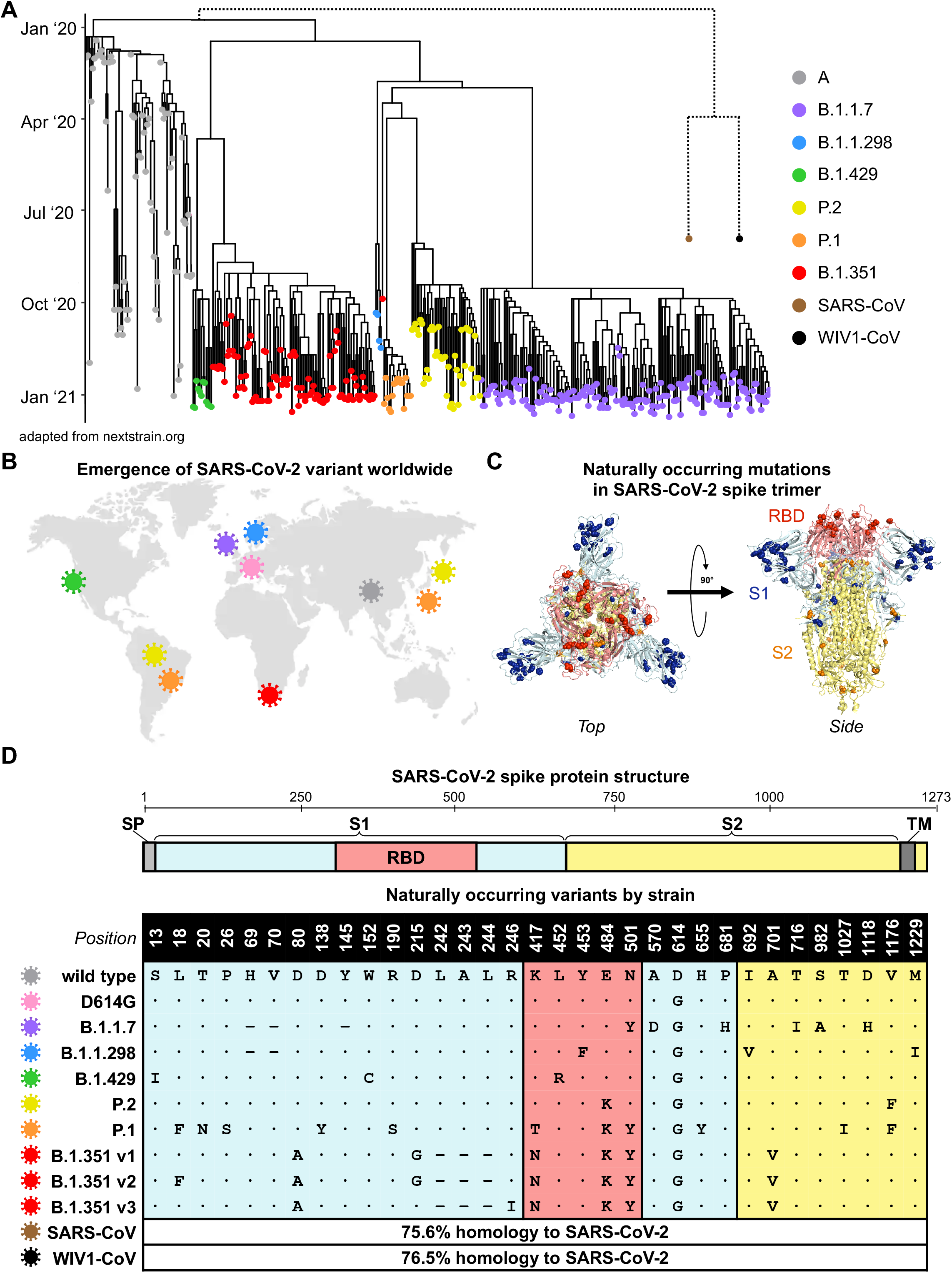
Emergence and spread of SARS-CoV-2 variants of concern around the world. (**A**)Phylogenetic tree of SARS-CoV-2 variants (adapted from nextstrain.org) with sampling dates is illustrated with a focus on the following lineages: A (grey), B.1.1.7 (purple), B.1.1.298 (blue), B.1.429 (green), P.2 (yellow), P.1 (orange), and B.1.351 (red). Dotted lines to SARS-CoV (brown) and bat-derived WIV1-CoV (black) are not to scale but indicate a distant phylogenetic relationship to SARS-CoV-2. (**B**)World map depicting the locations where the variants of these lineages were first described: original wild-type virus from A lineage (grey) in Wuhan, China; D614G variant (pink) in Europe that became dominant circulating strain; B.1.1.7 lineage (purple) in the United Kingdom; B.1.1.298 (blue) in Denmark; B.1.429 (green) in California, United States; P.2 (yellow) in Brazil and Japan; (orange) in Brazil and Japan; and B.1.351 (red) in South Africa. (**C**)Crystal structure of pre-fusion stabilized SARS-Cov-2 spike trimer (PDB ID 7JJI) is shown with top (*left panel*) and side (*right panel*) views. Sites where naturally occurring mutations occur are indicated with residue atoms highlighted as colored spheres. Spike regions and associated mutations are colored as follows: receptor binding domain (RBD) in red, S1 (excluding RBD) in blue, and S2 in yellow. (**D**)Schematic of SARS-CoV-2 spike protein structure and the mutation landscape of variants used in this study are illustrated. The mutations present in each variant tested represent the consensus sequence for that lineage and represent actual circulating strains: A (wild type), B.1.1.7, B.1.1.298, B.1.1.429, P.2, and P.1 lineages. In the case of the B.1.351 lineage, the three most abundant variants (v1, v2, and v3) deposited in GISAID were assessed. For SARS-CoV and WIV1-CoV, the percent homology is indicated. The following abbreviations are used: SP, signal peptide; TM, transmembrane domain; RBD, receptor binding domain.In the mutation map, a dot (**·**) indicates the same amino acid in that position as wild type and a dash (**–**) indicates a deletion.

### Vaccine-elicited neutralizing responses to circulating SARS-CoV-2 strains

We accrued a cohort of 99 individuals who received either one or two full doses of the BNT162b2 or mRNA-1273 vaccines, and used a luminescence-based lentiviral pseudovirus neutralization assay that we and others have previously validated (C. Wang et al. 2020; Ju et al. 2020; Pinto et al. 2020; Yang et al. 2020; Moore et al. 2004; Crawford et al. 2020; Garcia-Beltran et al. 2021) to assess neutralization of SARS-CoV-2 variants in a high-throughput system (**Figure 2A-B**). This cohort was relatively young (median age: 33 years, range: 22 - 73 years) and was 62.6% (62/99) female and 37.4% (37/99) male (**Figure S2**). We compared sera from 1220 pre-pandemic samples we had previously analyzed using the same assay (Garcia-Beltran et al. 2021) to our vaccinee cohort. Receiver operating characteristic analysis showed that the pseudovirus neutralization assay was able to accurately discriminate between non-vaccinated individuals and vaccinees ≥7 days following the second dose of BNT162b2 or mRNA-1273 (AUC = 0.9998), with a sensitivity of 100% and specificity of 99% using the limit of detection of the assay (pNT50 of 12) as the cut-off (**Figure S3**). Between non-vaccinated and individuals receiving 1 dose of vaccine (or <7 days following the second dose), the sensitivity was reduced to 93% (**Figure S3**). Sera obtained from individuals receiving 2 full doses of vaccine exhibited robust neutralization of wild-type SARS-CoV-2 pseudovirus, with BNT162b2 recipients achieving a median titer of 2,016 and mRNA-1273 recipients achieving a median titer of 762 (this difference was not statistically significant) (**Figure 3A**). Individuals who received only one dose of vaccine (or <7 days following the second dose) had significantly lower, but readily detectable, neutralization with a median titers of 167 for BNT162b2 and 208 for mRNA-1273 (**Figure 3C**). When comparing pseudovirus neutralization titer as a function of time post-vaccination, we observed an expected increase in titer after the second dose (**Figure 3B**). The magnitude of neutralization correlated with total anti-RBD antibody levels; however, this correlation was poor given that most vaccinees reached the upper limit of detection of our ELISA (**Figure 3C**). Neither sex nor age appeared to significantly influence neutralization titers in our cohort (**Figure 3D-B**).

**Figure 2.**
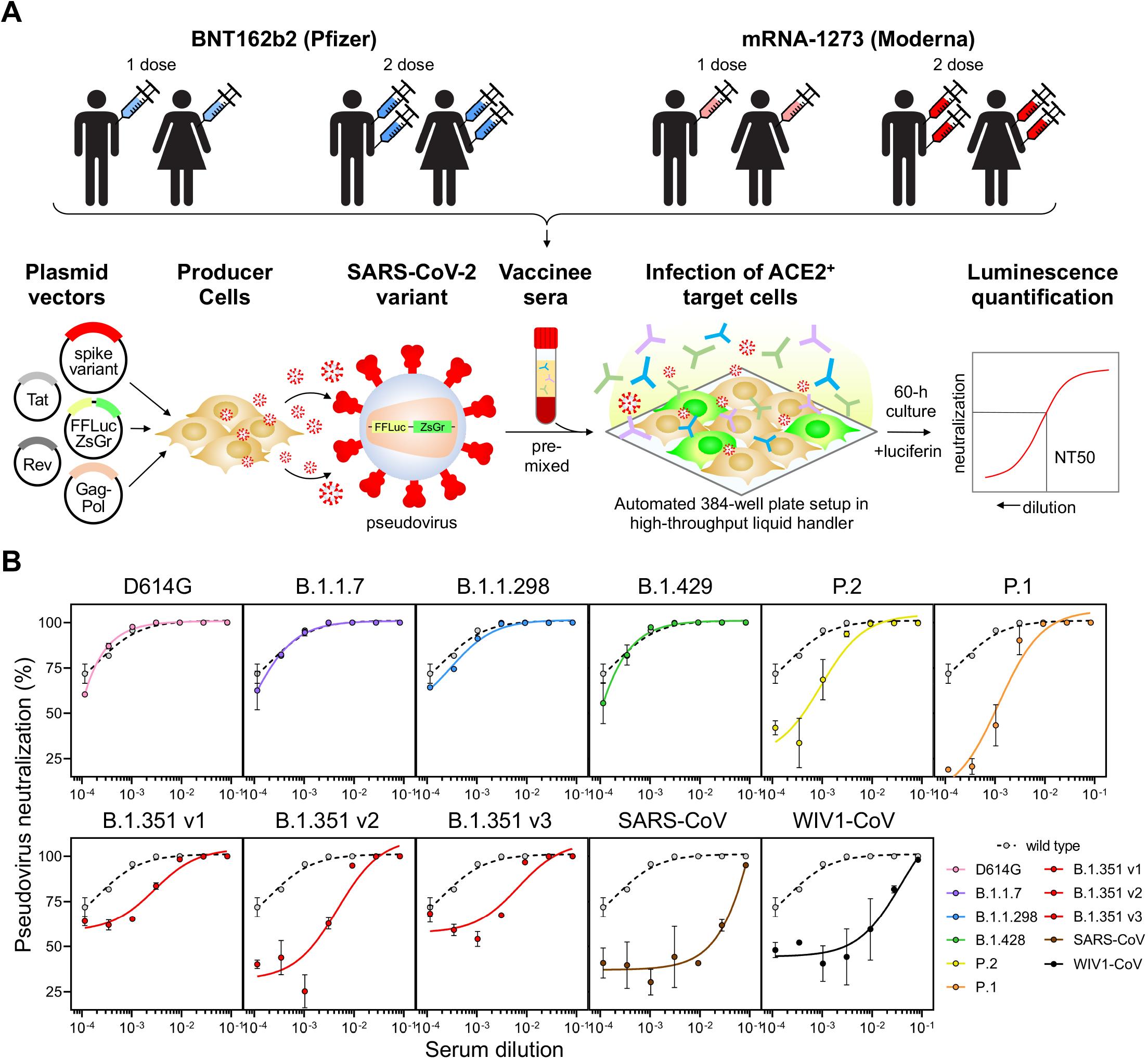
Neutralization of SARS-CoV-2 pseudovirus variants by vaccinee sera. (**A**)Schematic of our vaccine recipient cohort, consisting of individuals who received one or two full doses of either BNT162b2 (Pfizer) or mRNA-1273 (Moderna) vaccine, is presented in conjunction with our previously described high-throughput lentiviral vector-based SARS-CoV-2 pseudovirus neutralization assay (Garcia-Beltran et al. 2021). (**B**)Representative pseudovirus neutralization curves are shown for an individual ≥7 days out from the second dose of BNT162b2 vaccine comparing wild-type SARS-CoV-2 pseudovirus to the following variant pseudoviruses: D614G (pink); B.1.1.7 (purple); B.1.1.298 (blue); B.1.429 (green); P.2 (yellow); P.1 (orange); B.1.351 v1, v2, and v3 (red); SARS-CoV (brown); and WIV1-CoV (black).

**Figure 3.**
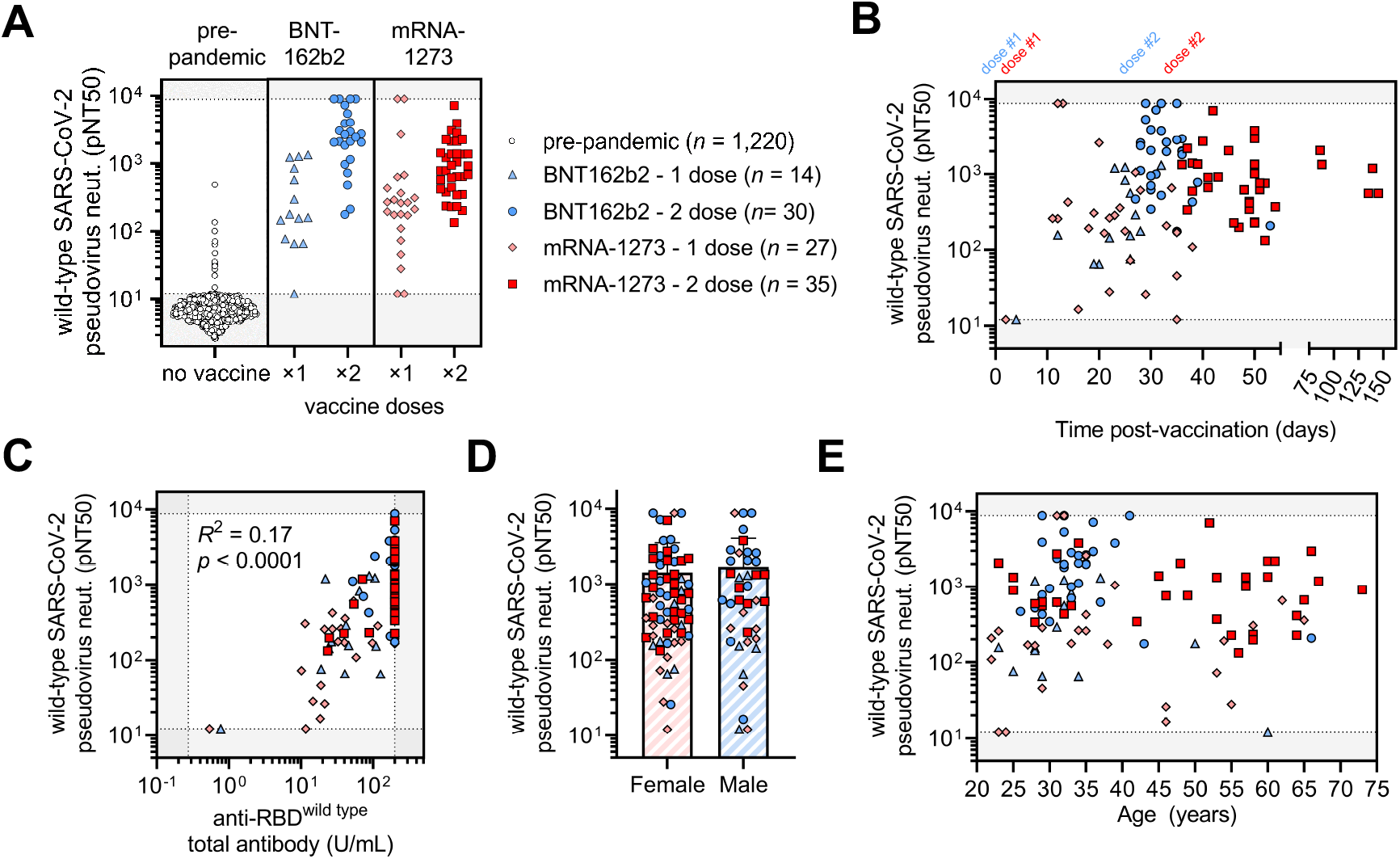
COVID-19 vaccines elicit potent but dose-dependent neutralizing responses against SARS-CoV-2 regardless of age or sex. (**A**)Titers that achieve 50% pseudovirus neutralization (pNT50) of wild-type SARS-CoV-2 are plotted for the following cohorts: (*i*) pre-pandemic individuals who were never vaccinated or infected with SARS-CoV-2 (pre-pandemic, white circles, *n* = 1,220); (*ii*) vaccine recipients that received only one dose of the BNT162b2 vaccine or were <7 days from their second dose (BNT162b2 - 1 dose, light blue triangles, *n* = 14); (*iii*) vaccine recipients ≥7 days following their second dose of BNT162b2 vaccine (BNT162b2 - 2 dose, blue circles, *n* = 30); (*iv*) vaccine recipients that received only one dose of the mRNA-1273 vaccine or were <7 days from their second dose (mRNA-1273 - 1 dose, pink diamonds, *n* = 27), and (*v*) vaccine recipients ≥7 days following their second dose of the mRNA-1273 vaccine (mRNA-1273 - 2 dose, red squares, *n* = 35). (**B**)Pseudovirus neutralization (pNT50) of wild-type SARS-CoV-2 is plotted versus time post-vaccination. The first dose (“dose #1”) of both BNT162b2 (blue) and mRNA-1273 (red) vaccines occurred on day 0. The second dose of the BNT162b2 vaccine occurred approximately 21 days after the first dose (“dose #2” in blue text) and for the mRNA-1273 vaccine occurred approximately 29 days after the first dose (“dose #2” in red text). Groups of vaccinees are the same as in (**A**). (**C**)Total anti-RBD (wild type) antibody levels measured by a quantitative ELISA against the wild-type RBD antigen versus neutralization of wild-type SARS-CoV-2 pseudovirus are presented. ELISAs were performed in duplicate and average values were used. Groups of vaccinees are the same as in (**A**). (**D**)Neutralization of wild-type SARS-CoV-2 pseudovirus is compared between female and male vaccinees. Bars and error bars indicate mean and standard deviation. Groups of vaccinees are the same as in (**A**). (**E**)Neutralization of wild-type SARS-CoV-2 pseudovirus is plotted against age of vaccine recipient. Groups of vaccinees are the same as in (**A**).

Neutralization of D614G pseudovirus was similar to that of wild type in individuals who received two doses of vaccine (1.2-fold decrease for both 2-dose vaccines) (**Figure 4A-B** and **S4A**), which was in contrast to previous studies in convalescent sera that we and others conducted demonstrating slightly increased neutralization of D614G variant versus wild type (Garcia-Beltran et al. 2021) following natural infection. The effect was more pronounced in individuals who received one dose of vaccine, some of which had undetectable neutralization of D614G despite detectable neutralization of wild-type SARS-CoV-2 (**Figure 4A-B**). This difference may be a consequence of the vaccine encoding the wild-type spike sequence, while many convalescent individuals in previous studies were likely infected with D614G variant SARS-CoV-2, given it had already become the globally dominant strain by the summer of 2020 (Lemieux et al. 2020; Korber et al. 2020).

**Figure 4.**
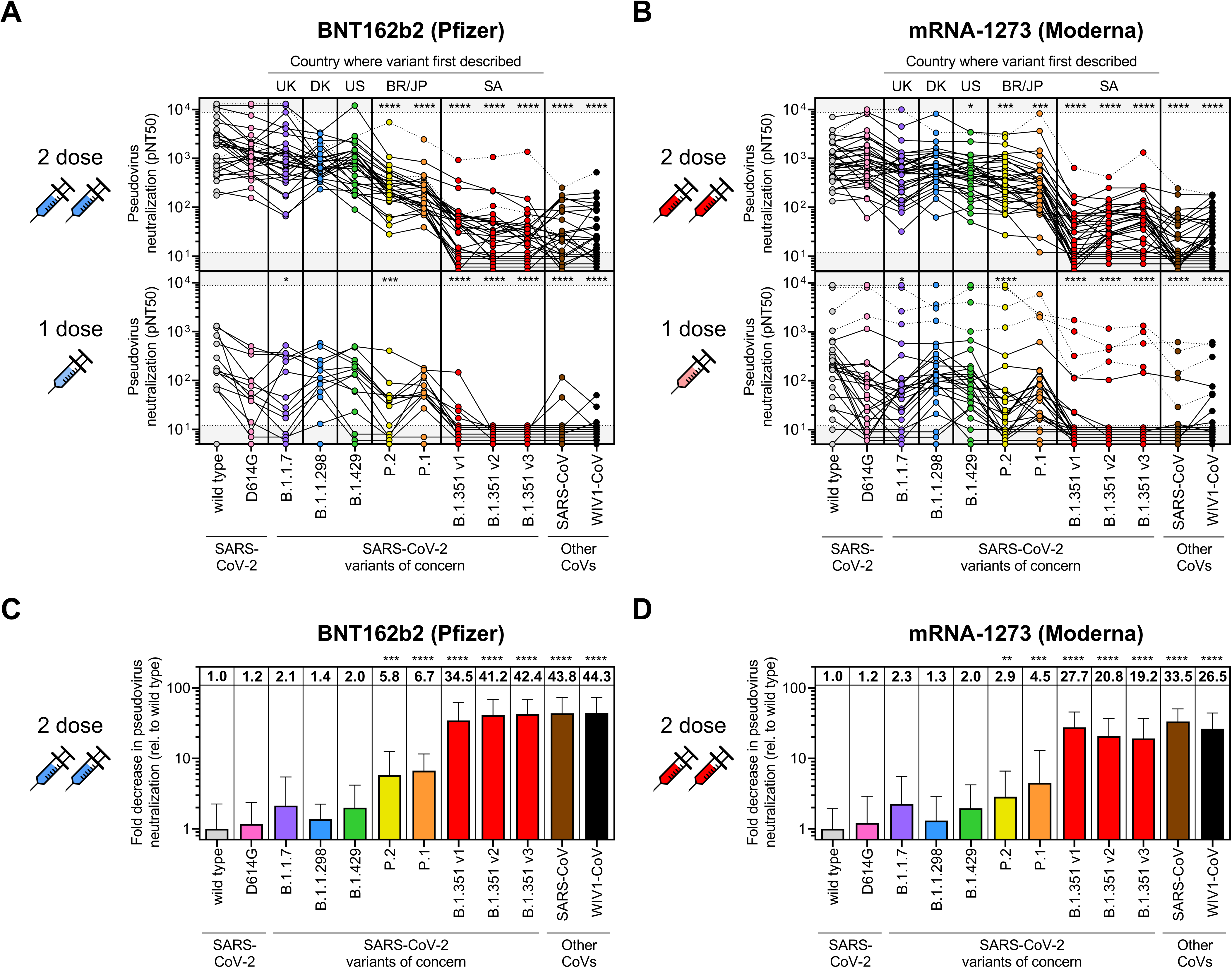
Sera from COVID-19 vaccine recipients cross-neutralize some but not all SARS-CoV-2 variants of concern. (**A**-**B**) Pseudovirus neutralization (pNT50) is plotted for all individuals that received 1 dose (*bottom panels*) or 2 full doses (*upper panels*) of either the BNT162b2 (**A**) or mRNA-1273 (**B**) vaccines for each of the following SARS-CoV-2 pseudoviruses: wild-type; D614G; B.1.1.7; B.1.1.298; B.1.429; P.2; P.1; and three variants of the B.1.351 lineage denoted as B.1.351 v1, v2, and v3. Other distantly related coronaviruses, namely, SARS-CoV from the 2002 Hong Kong outbreak and the pre-emergent bat coronavirus WIV1-CoV, were also tested. Individuals who were <7 days out from their second dose of the vaccine were classified as having received 1 dose. Dotted lines indicate vaccine recipients that were previously diagnosed with or highly suspected to have COVID-19 before being vaccinated. Gray regions indicate upper and lower limits of detection of our neutralization assay. The following abbreviations are used to indicate the country where the variant was first described: UK, United Kingdom; DK; Denmark; US, United States; BR, Brazil; JP, Japan; and SA, South Africa. An ANOVA correcting for multiple comparisons was performed and statistical significance of each pseudovirus relative to wild type is shown with the following notations: * *p* < 0.05, *** *p* < 0.001, **** *p* < 0.0001. (**C-D**) Fold decrease in neutralization for each pseudovirus relative to wild type is shown for vaccine recipients ≥7 days out from the second dose of BNT162b2 (*n* = 30) (**A**) or mRNA-1273 (*n* = 35) (**B**). Fold decrease was calculated by dividing the concentration at which 50% neutralization is achieved (IC50, which is 1/NT50) by the average IC50 value of wild type. The value of the mean is shown at the top of each bar. Bars and error bars indicate mean and standard deviation. An ANOVA correcting for multiple comparisons was performed and statistical significance of each pseudovirus relative to wild type is shown with the following notations: ** *p* < 0.01, *** *p* < 0.001, **** *p* < 0.0001.

When assessing variants containing one RBD mutation as part of their mutational landscape, the UK variant B.1.1.7 (N501Y), Danish mink variant B.1.1.298 (Y453F), and California variant B.1.429 (L452R) exhibited neutralization that was generally similar to that of wild-type and the parental D614G variant. Among people who received 2 full doses of BNT162b2, the mean fold decrease in neutralization relative to wild type was 2.1-fold for B.1.1.7, 1.4-fold for B.1.1.298, and 2.0-fold for B.1.429 (**Figure 4C** and **S4A**); for those who received 2 full doses of mRNA-1273, the mean fold decrease in neutralization relative to wild type was 2.3-fold for B.1.1.7, 1.3-fold for B.1.1.298, and 2.0-fold for B.1.429 (**Figure 4D** and **S4A**). However, neutralization of the Brazilian/Japanese P.2 variant, whose RBD contains an E484K mutation, was significantly decreased (5.8-fold for BNT162b2, *p* < 0.001; 2.9-fold for mRNA-1273, *p* < 0.01) (**Figure 4C-B** and **S4A**). This is in line with previous studies suggesting that the E484K mutation can evade polyclonal antibody responses (Greaney et al. 2020; Jangra et al. 2021) and has been found in cases of SARS-CoV-2 re-infection (Paiva et al. 2020; Faria et al. 2021; Resende et al. 2021; Naveca et al. 2021; Nonaka et al. 2021). Similarly, neutralizing antibody responses were also significantly decreased for the Brazilian/Japanese P.1 strain (6.7-fold for BNT162b2, *p* < 0.0001; 4.5-fold for mRNA-1273, *p* < 0.001), which harbors three mutations in RBD (K417T, E484K, and N501Y) and has also been found in cases of re-infection (nuno_faria 2021).

Strikingly, neutralization of all three South African B.1.351 strains was substantially decreased for both 2-dose vaccines (v1: 34.5-fold for BNT162b2 and 27.7-fold for mRNA-1273; v2: 41.2-fold for BNT162b2 and 20.8-fold for mRNA-1273; v3: 42.4-fold for BNT162b2 and 19.2-fold for mRNA-1273; *p* < 0.0001 for all comparisons) (**Figure 4C-B** and **S4A**). These strains contain the same three RBD mutations as P.1 except for an asparagine versus threonine substitution at K417 (K417N) and several additional mutations in non-RBD regions. Surprisingly, we found that the decreased in neutralization of B.1.351 strains was similar to that of distantly related coronaviruses, namely, SARS-CoV (43.8-fold for BNT162b2 and 33.5-fold for mRNA-1273) and WIV1-CoV (44.3-fold for BNT162b2 and 26.5-fold for mRNA-1273), suggesting that a relatively small number of mutations can medicate potent escape from vaccine-induced neutralizing antibody responses. These substantial differences in neutralization could not be explained by differences in spike protein expression (**Figure S4B**) nor in the amount of pseudovirus used in the assay (**Figure S4C**). Notably, 36.7% (11/30) recipients of 2-dose BNT162b2 and 42.9% (15/35) recipients of 2-dose mRNA-1273 vaccines did not have detectable neutralization of at least one of the B.1.351 variants (**Figure 4A-B**). Of the individuals who received only 1 dose of BNT162b2 or mRNA-1273, all of them had undetectable neutralization of B.1.351 v2 and v3, except for four individuals who reported having prior COVID-19 infection or a significant exposure and one individual for which a COVID-19 history was not obtained. This suggests that a single dose of existing mRNA vaccines may be insufficient to induce cross-neutralizing antibody responses in previously uninfected individuals. Interestingly, the six individuals in our study who reported having prior COVID-19 infection or significant exposure had among the highest neutralization titers for most variants and exhibited substantial cross-neutralization of the B.1.351, SARS-CoV, and WIV1-CoV (**Figure 4A-B** and **S2**). This suggests that prior infection combined with vaccination may result in the greatest breadth of cross-reactive neutralizing antibody responses, even against distantly related coronaviruses.

### Neutralization resistance of B.1.351 variants is primarily due to RBD mutations

To better characterize the mutational context responsible for B.1.351 neutralization resistance, we explored the contribution of mutations located both within and outside of the RBD region of spike. Of note, many of the substitutions and deletions found in the B.1.351 lineage are in close structural proximity in the S1 domain (**Figure 5A**). However, 86.2% of available GISAID sequences of this lineage harbor these mutations in three distinct patterns represented by B.1.351 v1 (46.6%), v2 (31.6%) and v3 (8.0%). We created pseudoviruses in which the RBD regions of B.1.351 v1, v2, and v3 were reverted to their original wild-type Wuhan sequence (v1/wtRBD, v2/wtRBD, and v3/wtRBD) while retaining all other B.1.351 v1, v2, or v3 mutations. Remarkably, neutralization assays conducted with sera from 24 individuals that received the 2-dose BNT162b2 vaccine revealed that neutralization of B.1.351 v1, v2, and v3 in the absence of RBD mutations was comparable to that of D614G (**Figure 5B-B**). A pseudovirus bearing only the three RBD mutations (K417N, E484K, and N501Y), largely, but not entirely, recapitulated the escape phenotype (**Figure 5B** and **5D**). Despite this escape, antibodies exhibited reduced, but detectable, binding to mutant RBD protein harboring B.1.351 mutations (K417N, E484K, and N501Y) by ELISA, which correlated with K417N+E484K+N501Y pseudovirus neutralization (*R*^2^ = 0.67, *p* < 0.0001) (**Figure 5E**). This suggests that vaccine-elicited antibodies capable of binding mutant RBD can retain the ability to neutralize RBD-mutant pseudoviruses.

**Figure 5.**
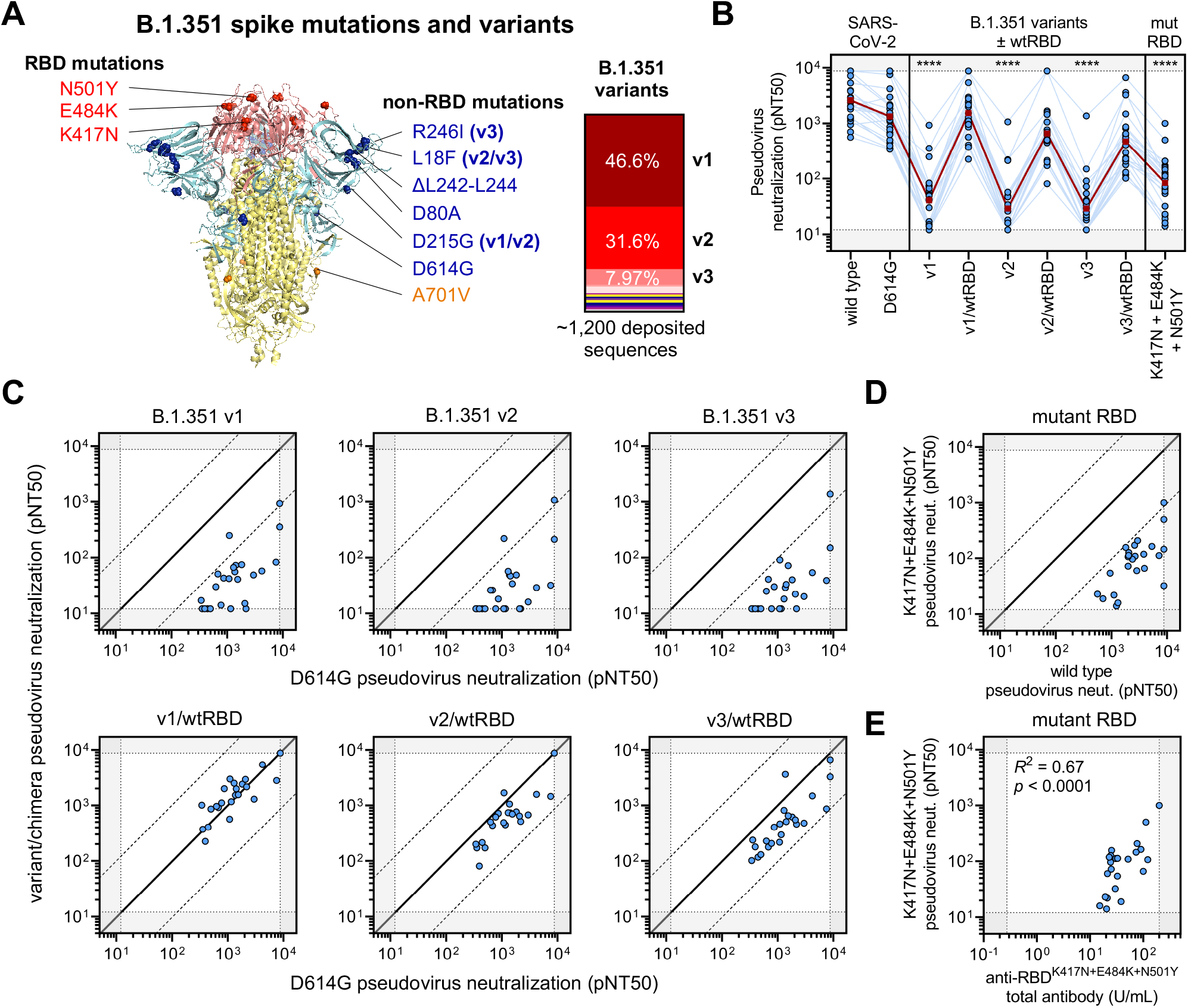
Limited cross-neutralization of B.1.351 strains of SARS-CoV-2 is mainly due to RBD mutations. (**A**)Crystal structure of pre-fusion stabilized SARS-Cov-2 spike trimer (PDB ID 7JJI) with RBD and non-RBD mutation sites for B.1.351 variants are indicated with residue atoms highlighted as colored spheres. Spike regions and associated mutations are colored as follows: RBD in red, S1 (excluding RBD) in blue, and S2 in yellow. In the case of mutations that are present in only some variants, the variants in which they occur (v1, v2, and/or v3) are indicated; all other mutations are present in the three tested B.1.351 variants. The relative frequencies of B.1.351 v1, v2, and v3 variants (as determined by sequences deposited in GISAID) are indicated (*right panel*). (**B**)Neutralization of B.1.351 v1, v2, and v3 variants was compared to chimeric variants lacking RBD mutations, denoted v1/wtRBD, v2/wtRBD, and v3/wtRBD, in 24 vaccine recipients ≥7 days out from the second dose of BNT162b2. Sera was also tested against pseudovirus bearing only RBD mutations found in B.1.351 (K417N, E484K, and N501Y). The dark red line indicates the geometric mean of the relative neutralization of each pseudovirus. Gray regions indicate upper and lower limits of detection of the assay. (**C**)Correlations between pseudovirus neutralization titer of D614G and B.1.351 chimeric viruses are shown. The solid diagonal line of identity indicates identical neutralization, and the dotted diagonal black lines indicate a 10-fold difference in pseudovirus neutralization (pNT50). (**D**)Pseudovirus neutralization (pNT50) of K417N+E484K+N501Y mutant pseudovirus is correlated to wild type pseudovirus. The dark red line indicates the geometric mean of the relative neutralization of each pseudovirus. Gray regions indicate upper and lower limits of detection of the assay. (**E**)Total antibodies that bind RBD harboring the B.1.351 mutations (anti-RBD^K417N+E484K+N501Y^ total antibodies) were measured by a quantitative ELISA and correlated to pseudovirus neutralization (pNT50) of K417N+E484K+N501Y mutant pseudovirus.

Upon assessing the contribution of RBD versus non-RBD mutations towards escape, regression analyses provided statistical evidence of interaction between mutations within and outside of RBD in mediating escape from B.1.351 variants, consistent with synergistic (i.e. non-additive) effects of these mutations (interaction statistics for B.1.351 v1 was *p* = 0.06, v2 was *p* < 0.0001, and v3 was *p* < 0.0001). These results suggest that while RBD mutations contribute the majority of the observed escape from vaccine-induced neutralization, they are more effective when in the context of additional mutations, particularly those found in B.1.351 v2 and v3 variants.

## DISCUSSION

Traditionally, polyclonal immune responses that arise in the context of infection and vaccination are thought to target multiple antigenic epitopes. Given this assumption, the expectation would be for small numbers of variations in antigen sequence to have only modest effects on recognition by the immune system. Here, we find that while many strains, such as B.1.1.7, B.1.1.298, or B.1.429, continue to be potently neutralized despite the presence of individual RBD mutations, other circulating SARS-CoV-2 variants escape vaccine-induced humoral immunity. The P.2. variant, which contains an E484K mutation within the RBD region, was capable of significantly reducing neutralization potency of fully vaccinated individuals, in line with what has been suggested by deep mutational scanning (Greaney et al. 2020; Jangra et al. 2021). Similarly, the P.1 strain, which has three RBD mutations, more effectively escaped neutralization, possibly explaining recently reported cases of re-infection with this variant (Paiva et al. 2020; Faria et al. 2021; Resende et al. 2021; Naveca et al. 2021; Nonaka et al. 2021). Finally, we found that B.1.351 variants exhibited remarkable resistance to neutralization, largely due to three mutations in RBD but with measurable contribution from non-RBD mutations. The magnitude of the effect is such that B.1.351 strains escaped neutralizing vaccine responses as effectively as distantly related coronaviruses.

Given the loss of vaccine potency against a number of circulating variants, most individuals receiving a single dose of vaccine did not raise sufficient antibody titers to provide any detectable cross neutralization against B.1.351 v2 or v3. While our studies are limited by the relatively short follow-up time after vaccination, our findings support the importance of 2-dose regimens to achieve titers, and perhaps breadth, to enhance protection against novel variants. These findings are important to consider in the context of proposals to administer a single dose of vaccine across a larger number of individuals instead of using doses to boost prior recipients

Importantly, our studies rely on pseudoviruses which are only capable of modeling the ACE2-dependent entry step of the SARS-CoV-2 lifecycle. While numerous studies have now demonstrated a close correlation between neutralization titers measured against pseudovirus and live SARS-CoV-2 cultures (C. Wang et al. 2020; Ju et al. 2020; Pinto et al. 2020; Yang et al. 2020; Moore et al. 2004; Crawford et al. 2020; Riepler et al. 2020), it is unclear what impact additional mutations located outside of the spike may have on immunological escape, virulence, infectivity, or pathogenesis. Several recent reports and pre-prints, including studies conducted by Pfizer as well as Moderna, have produced similar findings in terms of vaccine potency against B.1.1.7 and B.1.1.298 variants, but substantially less neutralization resistance by B.1.351 than we measured. However, we would caution that each study was done with distinct serum samples measured using different conditions (e.g. replication-competent chimeric vesicular stomatitis virus versus single-entry lentiviral pseudovirus) and with different spike expression plasmids and combinations of mutations (e.g. triple RBD mutations, lineage-defining mutations, full complement of mutations not yet observed together in nature, etc.). In the present study, we mimicked the natural pattern of mutations found in circulating strains, in phase. Importantly, these include the spike variants found in two replication-competent live-virus strains of the B.1.351 lineage, which were recently reported to exhibit nearly complete neutralization resistance in response to convalescent plasma (Cele et al. 2021). The strains used in the aforementioned study were designated as 501Y.V2.HVdF002 and 501Y.V2.HV001, which have identical spike protein sequences as the B.1.351 v1 and B.1.351 v2 pseudoviruses reported in this study.

One important aspect of immunity not addressed by our work is cellular immunity contributed by cytotoxic lymphocytes, including T and NK cells. Even in the absence of neutralizing humoral immunity, previous studies have suggested that cellular immunity can mitigate severe or prolonged infection (Le Bert et al. 2020). In convalescent individuals, T-cell immunity would not be restricted to spike-derived epitopes, but also from other more abundant proteins such as nucleocapsid. As such, it would be reasonable to assume that T-cell-mediated immunity elicited by infection would remain largely intact for circulating variants including B.1.351. Indeed, although recent studies by Johnson & Johnson have demonstrated reduced overall efficacy in South Africa, there was substantially more protection against severe or fatal disease than for mild-to-moderate disease (Herper et al. 2021). However, with the exception of killed whole virus vaccines, all currently available vaccine designs only provide spike protein as the target immunogen, thus limiting T-cell immunity to spike epitopes. Notwithstanding, one recent study has demonstrated that mutations in spike epitopes do not impair T-cell responses despite escaping neutralizing antibodies (Skelly et al. 2021).

In summary, our data highlights the challenges facing all vaccines whose designs were finalized early in the pandemic and based on the sequence of the first-reported virus from Wuhan, China. Given the global scale and magnitude of the ongoing pandemic, including case reports of re-infection, it is clear that viral evolution will continue. It is possible that current vaccines will still provide clinical benefit against variants that exhibit poor cross-neutralization, such as P.1 and B.1.351, by reducing COVID-19 disease severity, but this has yet to be determined. Ultimately, it will be important to develop interventions capable of preventing transmission of diverse SARS-CoV-2 variants, including vaccine boosters that target these variants or technologies capable of eliciting or delivering broadly neutralizing antibodies.

## LIMITATIONS OF THE STUDY

The main focus of this study was to assess the potential of vaccinee sera to neutralize circulating SARS-CoV-2 variants. For this, we developed a high-throughput lentiviral pseudovirus-based *in vitro* neutralization assay that uses an engineered 293T-ACE2 cell line as target cells. As such, these cells restrict pseudovirus entry in an ACE2-dependent manner and lack other cell-surface proteins that may play a role in natural infection, such as TMPRSS2 (Hoffmann et al. 2020) or NRP1 (Cantuti-Castelvetri et al. 2020). Additional studies are needed to assess the influence of these proteins on the serum neutralization titers measured. We did not assess other antibody-mediated functions such as complement deposition, antibody-dependent cellular cytotoxicity, or antibody-dependent cellular phagocytosis, which may contribute to protection even in the absence of neutralizing antibodies. In addition, we did not assess the role of vaccine-elicited cellular immune responses mediated by T cells and NK cells, which are likely to play a key role in disease prevention for vaccine recipients.

## Data Availability

This study assessed the neutralization potency of sera obtained from vaccinated individuals and was not a clinical trial. De-identified raw data of the results of serological assays can be provided upon request.

## ACKNOWLEDGEMENTS, FUNDING SUPPORT

We wish to thank Michael Farzan, PhD for providing ACE2-expressing 293T cells. We also thank Mandakolathur Murali, MD for many useful and insightful discussions. B.M.H. and is supported by award Number T32GM007753 from the National Institute of General Medical Sciences. J.F. is supported by T32AI007245. D.J.G., M.C.P., and M.N.P. were supported by the VIC Innovation fund. A.S. was supported by the Bill and Melinda Gates Investment INV-018944 (AS) and by the South African Medical Research Council and the Department of Science and Innovation (TdO), A.G.S. was supported by NIH R01 AI146779 and a Massachusetts Consortium on Pathogenesis Readiness (MassCPR) grant. A.J.I. is supported by the Lambertus Family Foundation. A.B.B. was supported by the National Institutes for Drug Abuse (NIDA) Avenir New Innovator Award DP2DA040254, the MGH Transformative Scholars Program as well as funding from the Charles H. Hood Foundation. This independent research was supported by the Gilead Sciences Research Scholars Program in HIV.

## AUTHOR CONTRIBUTIONS

W.F.G.B., E.C.L., K.S.D., and A.B.B. designed the experiments. W.F.G.B., E.C.L., K.S.D., Z.G., A.D.N., V.N., and A.B.B. carried out experiments and analyzed data. J.F., B.M.H., and A.G.S. provided key reagents and useful discussions and insights. D.J.G., M.C.P., and M.N.P. provided human samples for the study. A.D.N. and V.N. contributed to statistical and sequence analyses. A.J.I. and V.N. provided key discussions and input into experimental design. W.F.G.B., V.N., and A.B.B. wrote the paper with contributions from all authors.

## SUPPLEMENTAL FIGURE TITLES AND LEGENDS

**Figure S1.**
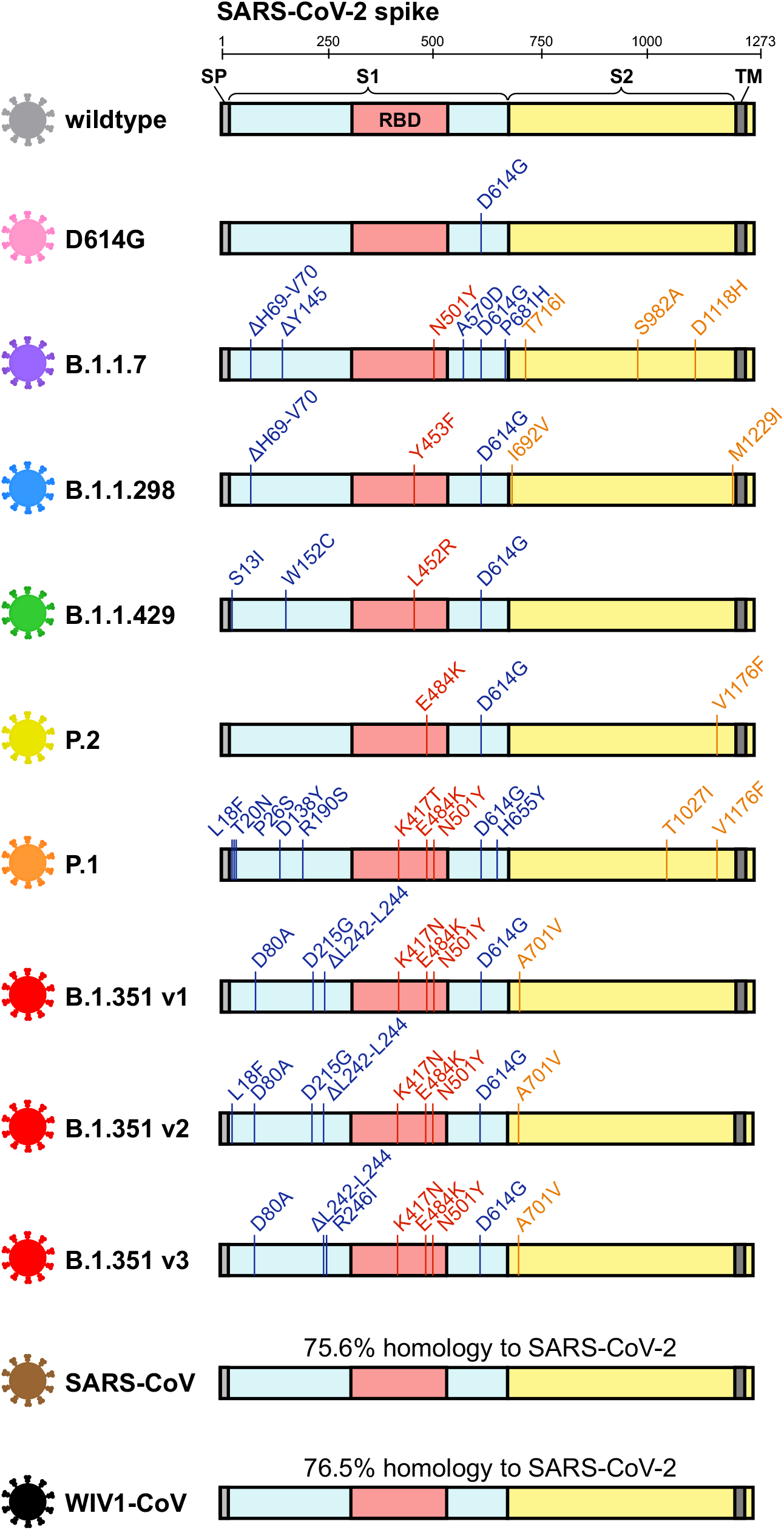
SARS-CoV-2 variants tested in this study, related to Figure 1. Schematic of mutations in the spike protein sequence of the following SARS-CoV-2 variants are illustrated: wild type (grey), D614G (pink), B.1.1.7 (purple), B.1.1.298 (blue), B.1.1.429 (green), (yellow), P.1 (orange), three variants of B.1.351 (red; v1, v2, and v3), SARS-CoV (brown), and WIV1-CoV (black).

**Figure S2.**
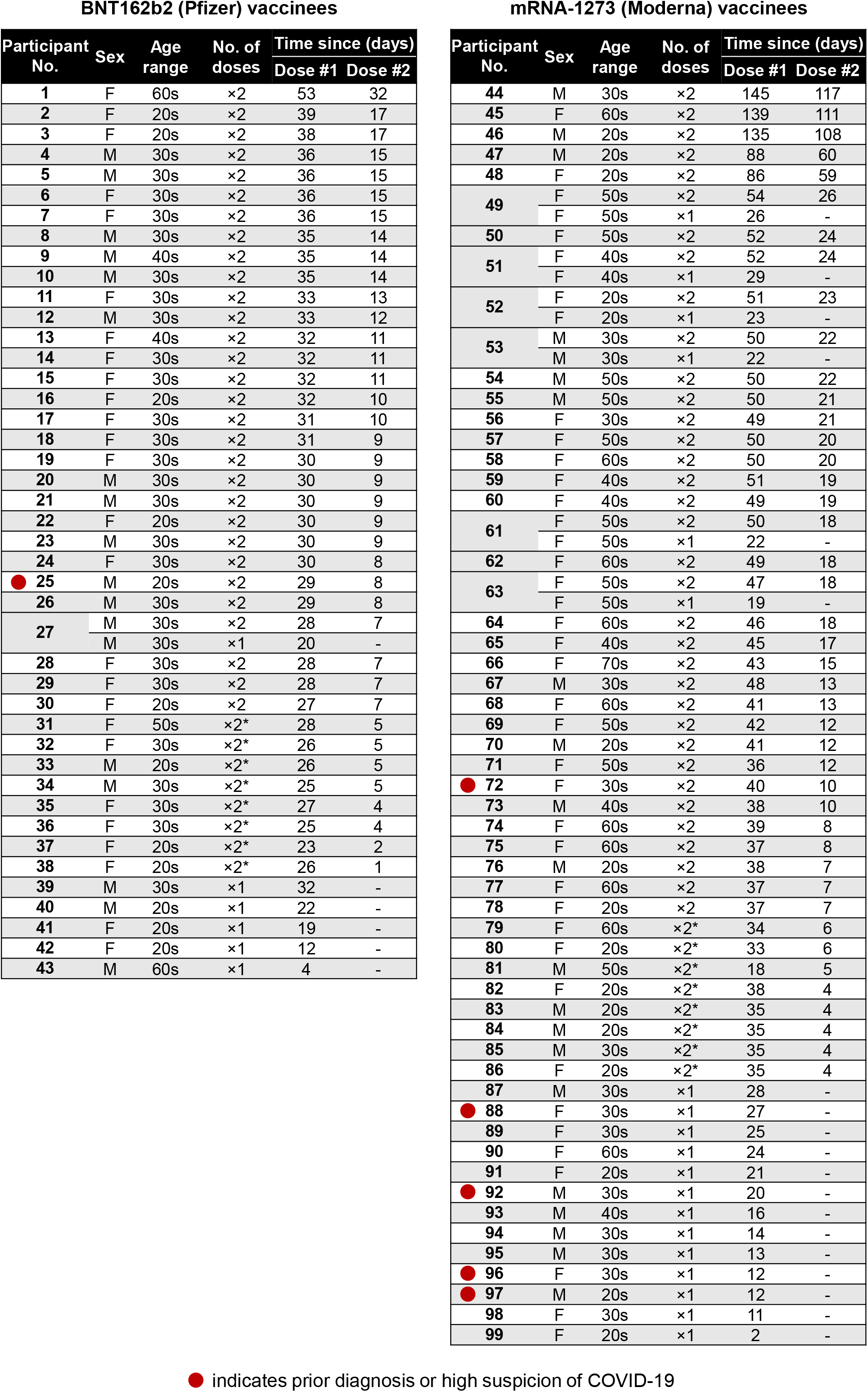
Characteristics of vaccine recipients, related to Figure 2. The sex and age of 99 vaccine recipients are presented along with time (in days) after the first and, when applicable, second doses of either the BNT162b2 (*left panel*) or mRNA-1273 (*right panel*) vaccines. Individuals who reported having been diagnosed with or highly suspected to have COVID-19 before vaccination are highlighted with a red circle.For 7 out of the 99 vaccinees, two samples taken, one after the first dose and one after the second dose.

**Figure S3.**
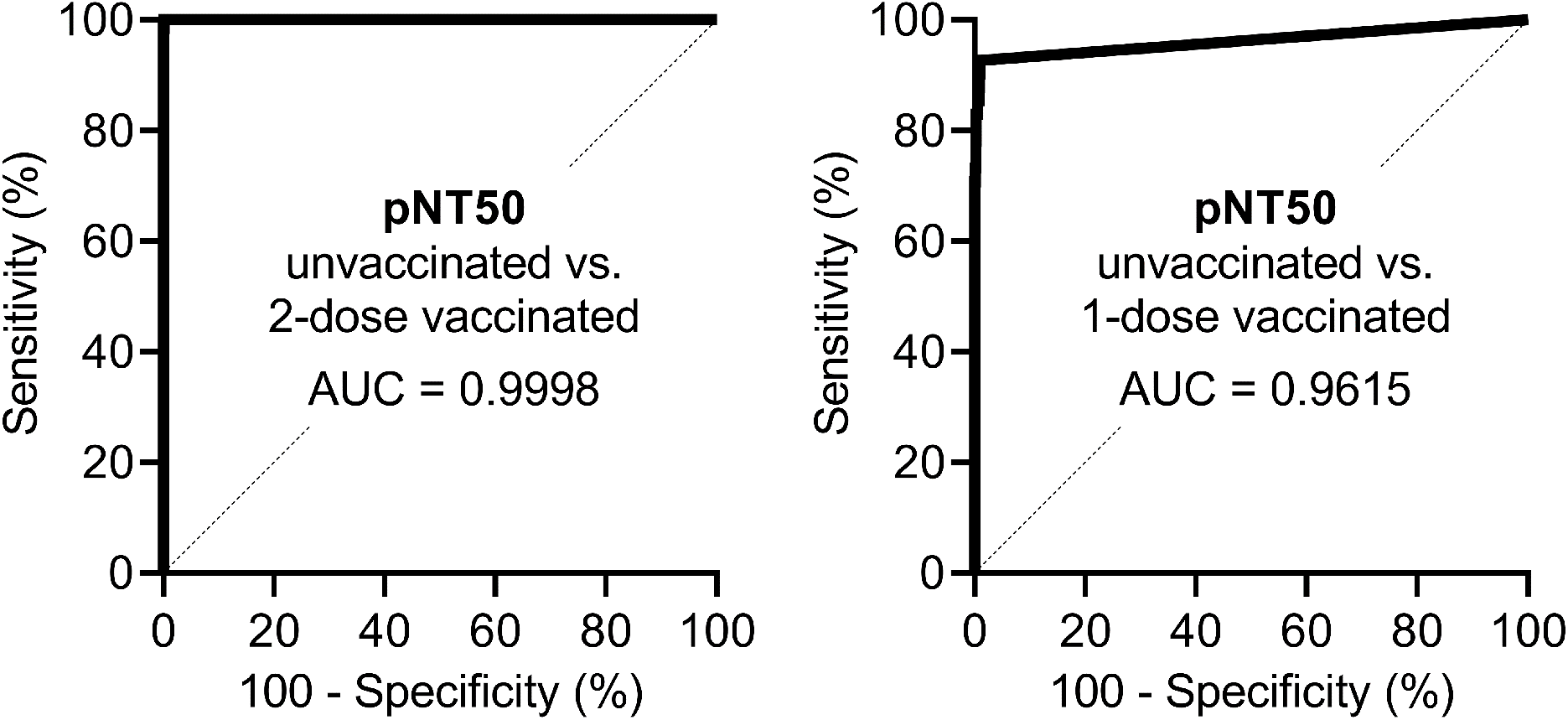
Receiver operating characteristic analysis of pseudovirus neutralization assay in vaccinated and non-vaccinated individuals, related to Figure 3. (**A-B**) Receiver operating curve (ROC) analysis was performed in non-vaccinated pre-pandemic individuals (*n* = 1,220) and recipients of 2 full doses of either BNT162b2 or mRNA-1273 (2-dose vaccinated, *n* = 65) (**A**) or 1 dose (or <7 days from second dose) of either vaccine (1-dose vaccinated, *n* = 41) (**B**). Area under the curve (AUC) is presented.

**Figure S4.**
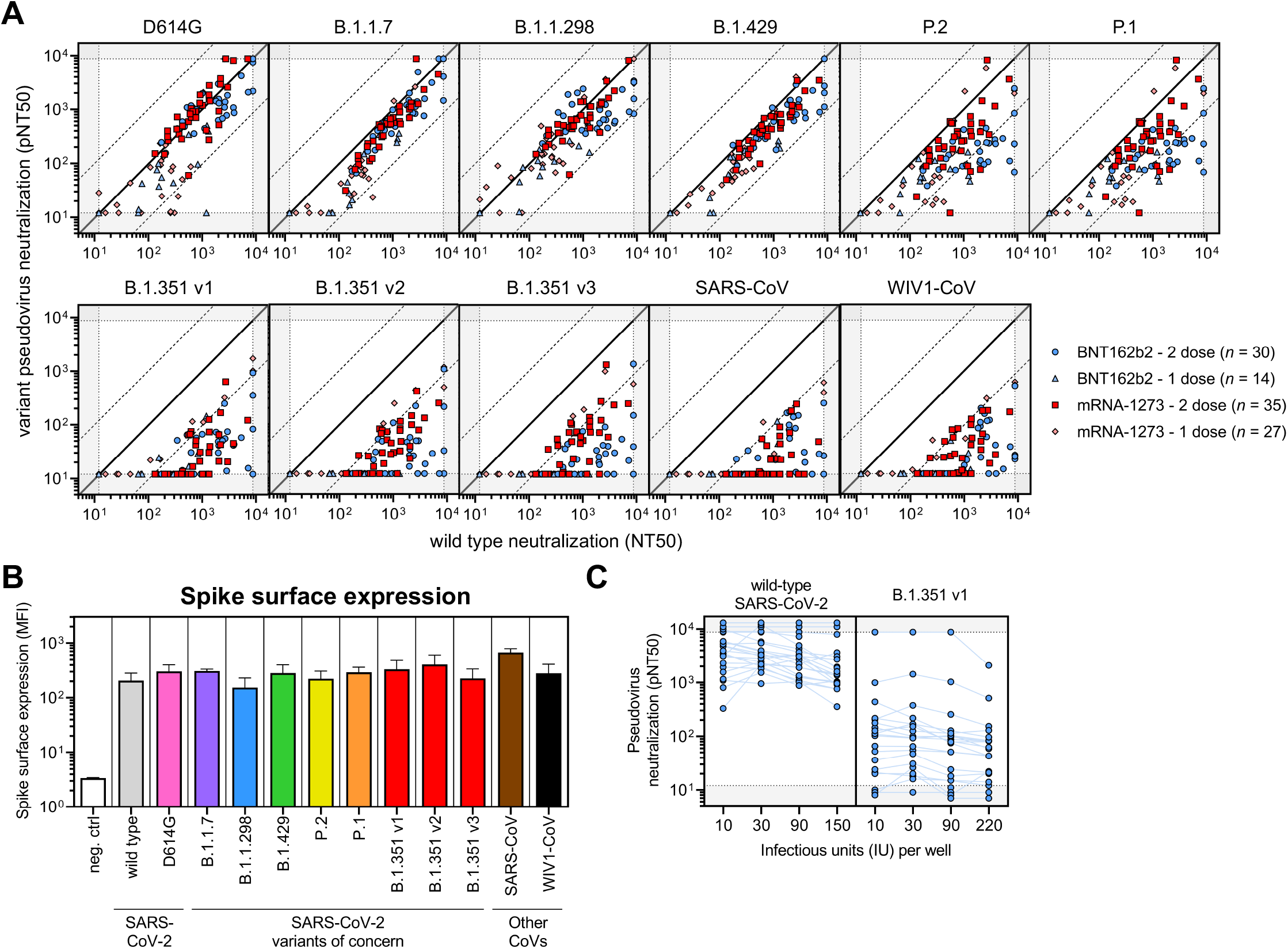
Correlations of SARS-CoV-2 variant neutralization titers, spike surface expression, and neutralization titers over a wide range of pseudovirus quantities, related to Figure 4. (**A**)Correlations between neutralization of wild-type and the indicated variant pseudoviruses are demonstrated. The solid diagonal black line indicates identical neutralization, and the dotted diagonal black lines indicate a 10-fold difference in pseudovirus neutralization (pNT50). Four groups of vaccine recipients are indicated: (*i*) vaccine recipients ≥7 days out from their second dose of BNT162b2 vaccine (*n* = 22, blue circles); (*ii*) vaccine recipients that received only one dose of the BNT162b2 vaccine or were <7 days from their second dose (*n* = 7, light blue triangles); (*iii*) vaccine recipients ≥7 days out from their second dose of the mRNA-1273 vaccine (*n* = 4, red squares); and (*iv*) vaccine recipients that received only one dose of the mRNA-1273 vaccine or were <7 days from their second dose (*n* = 14, pink diamonds). (**B**)Surface expression of the indicated variant spike proteins on the surface of transfected 293T cells was measured by flow cytometry. Transfected cells were stained with three monoclonal antibodies targeting spike, S309, ADI-55689, and ADI-56046, and median fluorescence intensities (MFI) of transfected (GFP+) cells were averaged to obtain a relative MFI demonstrating efficient expression of all spikes at the cell surface. Bars and error bars indicate mean and standard deviation. (**C**)To determine the consistency of pseudovirus neutralization titers (pNT50) measured by this assay, varying amounts of infectious units per well of wild-type SARS-CoV-2 and B.1.351 v1 pseudovirus were used to perform the neutralization assay for 22 serum samples from BNT162b2 vaccine recipients ≥7 days out from their second dose. These data demonstrate the robustness of calculating pNT50 across a 22-fold range of infectious units of pseudovirus.

## STAR ★ METHODS

### RESOURCE AVAILABILITY

#### Lead Contact

Further information and requests for resources and reagents should be directed to and will be fulfilled by Alejandro Balazs.

#### Materials Availability

Plasmids generated in this study will be available through Addgene. Recombinant proteins and antibodies are available from their respective sources.

#### Data and Code Availability

This study did not generate sequence data or code. Data generated in the current study (including ELISA and neutralization) have not been deposited in a public repository but are available from the corresponding author upon request.

### EXPERIMENTAL MODEL AND SUBJECT DETAILS

#### Human subjects

Use of human samples was approved by Partners Institutional Review Board (protocol 2020P002274). Serum samples from vaccine recipients that received one or two doses of the BNT162b2 or mRNA-1273 vaccine were collected in red top tubes.For each individual, basic demographic information including age and sex as well as any relevant COVID-19 history was obtained.

#### Cell lines

HEK 293T cells (ATCC) were cultured in DMEM (Corning) containing 10% fetal bovine serum (VWR), and penicillin/streptomycin (Corning) at 37°C/5% CO_2_. 293T-ACE2 cells were a gift from Michael Farzan (Scripps Florida) and Nir Hacohen (Broad Institute) and were cultured under the same conditions. Confirmation of ACE2 expression in 293T-ACE2 cells was done via flow cytometry.

## METHOD DETAILS

### Construction of variant spike expression plasmids

To create variant spike expression plasmids, we performed multiple PCR fragment amplifications utilizing oligonucleotides containing each desired mutation (Integrated DNA Technology) and utilized overlapping fragment assembly to generate the full complement of mutations for each strain. Importantly we generate these mutations in the context of our previously described codon-optimized SARS-CoV-2 spike expression plasmid harboring a deletion of the C-terminal 18 amino acids that we previously demonstrated to result in higher pseudovirus titers. Assembled fragments were inserted into NotI/XbaI digested pTwist-CMV-BetaGlobin-WPRE-Neo vector utilizing the In-Fusion HD Cloning Kit (Takara). All resulting plasmid DNA utilized in the study was verified by whole-plasmid deep sequencing (Illumina) to confirm the presence of only the intended mutations.

### SARS-CoV-2 pseudovirus neutralization assay

To compare the neutralizing activity of vaccinee sera against coronaviruses, we produced lentiviral particles pseudotyped with different spike proteins as previously described (Garcia-Beltran et al. 2021). Briefly, pseudoviruses were produced in 293T cells by PEI transfection of a lentiviral backbone encoding CMV-Luciferase-IRES-ZsGreen as well as lentiviral helper plasmids and each spike variant expression plasmid. Following collection and filtering, production was quantified by titering via flow cytometry on 293T-ACE2 cells. Neutralization assays and readout were performed on a Fluent Automated Workstation (Tecan) liquid handler using 384-well plates (Grenier). Three-fold serial dilutions ranging from 1:12 to 1:8,748 were performed for each serum sample before adding 50–250 infectious units of pseudovirus for 1 h. Subsequently, 293T-ACE2 cells containing polybrene were added to each well and incubated at 37°C/5% CO_2_ for 60-72 h. Following transduction, cells were lysed using a luciferin-containing buffer (Siebring-van Olst et al. 2013) and shaken for 5 min prior to quantitation of luciferase expression within 1 h of buffer addition using a Spectramax L luminometer (Molecular Devices). Percent neutralization was determined by subtracting background luminescence measured in cell control wells (cells only) from sample wells and dividing by virus control wells (virus and cells only). Data was analyzed using Graphpad Prism and pNT50 values were calculated by taking the inverse of the 50% inhibitory concentration value for all samples with a pseudovirus neutralization value of 80% or higher at the highest concentration of serum.

### Pseudovirus titering

To determine the infectious units of pseudotyped lentiviral vectors, we plated 400,000 293T-ACE2 cells per well of a 12-well plate. 24 h later, three ten-fold serial dilutions of neat pseudovirus supernatant were made in 100 μL, which was then used to replace 100 μL of media on the plated cells. Cells were incubated for 48 h at 37°C/5% CO_2_ to allow for expression of ZsGreen reporter gene and harvested with Trypsin-EDTA (Corning). Cells were resuspended in PBS supplemented with 2% FBS (PBS+), and analyzed on a Stratedigm S1300Exi Flow Cytometer to determine the percentage of ZsGreen-expressing cells. Infectious units were calculated by determining the percentage of infected cells in wells exhibiting linear decreases in transduction and multiplying by the average number of cells per well determined at the initiation of the assay. At low MOI, each transduced ZsGreen cell was assumed to represent a single infectious unit.

### Flow cytometric assessment of spike expression

To compare the relative surface expression of pseudovirus spike variant proteins, we plated 400,000 293T cells per well of a 12-well plate.24 h later, 1 μg of variant spike expression plasmid and transfected using PEI. Cells were incubated for 48 h at 37°C and harvested into PBS+ cells transfected with each vector were divided and stained with 5 μg/mL of either S309, ADI-55689, or ADI-56046 for 30 minutes at room temperature. Cells were then washed with 1 mL PBS+, spun at 900 × *g*, and stained with 2 μg/mL of anti-human IgG-AF647 polyclonal antibody (Invitrogen) for 30 minutes at room temperature. Cells were washed with 1 mL of PBS+, spun at 900 × *g*, resuspended in 100 μL of PBS+, fixed with 100 μL 4% PFA and analyzed on a Stratedigm S1300Exi Flow Cytometer.

### SARS-CoV-2 receptor binding domain total antibody ELISA

Quantitative detection of total antibodies to SARS-CoV-2 receptor binding domain (RBD) was performed as previously described (Garcia-Beltran et al. 2021). Briefly, we used an indirect ELISA with a standard consisting of anti-SARS-CoV and -CoV-2 monoclonal antibody (CR3022) (IgG1 isotype). 96-well ELISA plates were coated with purified wild-type SARS-CoV-2 RBD. Plates were blocked with BSA and washed. A seven-point standard curve was created using CR3022-IgG1 starting at 2 μg/mL by performing 1:3 serial dilutions with dilution buffer, and serum samples were diluted 1:100 with dilution buffer. Diluted samples and standards were added to corresponding wells and incubated for 1 h at 37°C, followed by washing. Total antibodies were detected with anti-human IgG+IgA+IgM (H+L)-HRP (Bethyl) diluted 1:25,000 for a 30 min incubation at room temperature. After washing, TMB substrate (Inova) was added to each well and incubated for 5-15 min before stopping with 1 M H_2_SO_4_. Optical density (O.D.) was measured at 450 nm with subtraction of the O.D. at 570 nm as a reference wavelength on a SpectraMax ABS microplate reader. Anti-RBD antibody levels were calculated by interpolating onto the standard curve and correcting for sample dilution; one unit per mL (U/mL) was defined as the equivalent reactivity seen by 1 μg/mL of CR3022. In assays where mutated RBD harboring K417N, E484K, and N501Y mutations was used, the exact same ELISA protocol was performed after confirming that our standard (CR3022-IgG1) bound to mutant versus wild-type RBD nearly identically.

### QUANTIFICATION AND STATISTICAL ANALYSIS

Data and statistical analyses were performed using GraphPad Prism 9.0.1, JMP Pro 15.0.0 (SAS Institute), and R v4.0.2. Flow cytometry data was analyzed using FlowJo 10.7.1. Non-parametric multivariate ANOVAs were performed on the indicated figures where several cohorts were present; all *p* values were adjusted for multiple comparisons. Statistical significance was defined as *p* < 0.05. Error bars throughout all figures represent one standard deviation. Interaction/regression analyses were performed using the lm package in R, using pNT50 estimates (normalized to wild type) for each donor sera and allowing for interaction between neutralization estimates for each wild-type RBD variant (v1/wtRBD, v2/wtRBD, or v3/wtRBD) and the variant containing only K417N+E484K+N501Y mutations to explain the composite neutralization of B.1351 v1, v2, and v3.

